# CCOFEE-GI Study: Colombian COVID19 First Experience in Gastroentrology. Characterization of digestive manifestations in patients diagnosed with COVID-19 at a highly complex institution in Bogota D.C., Colombia

**DOI:** 10.1101/2020.07.24.20161604

**Authors:** Alejandro Concha Mejía, Reinaldo Rincón Sánchez, Douvan Calderón-Zapata, Santiago Huertas Tafur, Andrés Montoya Durana, John Guzmán Camacho, Alan Ovalle Hernández, Heinz Ibañez Varela, Juan Pablo Osorio Lombana, Edwin Silva Monsalve, Fabian Cortés-Muñoz

## Abstract

The current pandemic caused by SARS-CoV-2 has posed an important threat to the human health, healthcare systems, economy, and structure of societies. In Colombia, the first case was diagnosed on March 6, 2020, with exponential progressive growth, and there were >200,000 confirmed cases as of July 20, 2020, in this cross-sectional, analytical, and observational study, we focused on the demographic, epidemiologic, and clinical characteristics of patients with confirmed SARS-CoV-2 infection at a highly complex institution in Latinamerica, with special emphasis on gastrointestinal symptoms. Methods: Demographic and clinical data were collected, results related to the outcomes such as hospitalization time, admission to ICU, need for orotracheal intubation, and death were also included. Statistical analyses were conducted using Stata software V.15.

**Results:** We included 72 patients RT-PCR positive for SARS-CoV-2 (34 women and 38 men) with age 47.5 ± 17.7 years; 17 (23.6%) presented at least one of the gastrointestinal symptoms (nausea/vomiting, abdominal pain, and/or diarrhea). 13 (76.47%) presented with diarrhea, 29.41% with nausea/vomiting, and five (29.41%) with abdominal pain. Diarrhea in 18.06% of all those infected with SARS-CoV-2 at the time of consultation, which was the most common digestive symptom. No significant differences were observed in requirement for endotracheal intubation, hospitalization, ICU admission, and fatal outcome between the NGIS and GIS groups (p:0.671, 0.483, 1,000, and 1,000). Conclusion: In our study, patients with gastrointestinal symptoms had no significant differences in disease severity, admission to ICU or death compared to those who did not have such symptoms.

## INTRODUCTION

The current pandemic caused by severe acute respiratory syndrome 2 (SARS-CoV-2) has posed an important threat to the human health, healthcare systems, economy, and structure of societies. In Colombia, the first case was diagnosed on March 6, 2020 (1), with exponential progressive growth, and there were >200,000 confirmed cases as of July 20, 2020, almost 4 months later.

The initially recognized manifestations include fever, myalgia, cough, and fatigue. The most frequently described radiological findings correspond to predominantly interstitial pneumonia (2,3). The presences of anosmia and dysgeusia have also been described as the likely neurological symptoms of SARS-CoV-2 infection (4). This simple-chain RNA virus, a member of the family Coronaviridae, genus *Betacoronavirus*, has been found in rectal swabs and samples of fecal matter where they can remain for >30 days (5), suggesting that the virus can infect the digestive tract and replicate there (3,6). Studies have reported digestive symptoms in patients with COVID-19 such as anorexia 39.7%–50.2%, diarrhea 2%–49.5%, nausea 1%–10.1%, vomiting 1%–15.9%, and abdominal pain 0.96%–6% (2,7–13). Digestive bleeding has also been reported in inflammatory diarrhea secondary to colitis due to SARS-CoV-2 (14,15). Patients with digestive symptoms have a long time from the onset of symptoms to admission to hospital services, and the presence of these gastrointestinal manifestations is associated with a longer duration and severity of the clinical presentation of COVID-19 (16), with increased stay in the intensive care unit (10).

Some medicines used to treat COVID-19 may generate digestive symptoms as side effects such as lopinavir/ritonavir that can cause diarrhea, nausea, and abdominal pain or tocilizumab that is associated with an increase in transaminase level (9,17).

There is evidence for the expression of ACE2 and TMPRSS2 receptors in the gastrointestinal and hepatobiliary tracts, which may be a potential explanation for gastrointestinal and hepatic manifestations as well as a possible fecal–oral contamination route (2,8,9,18,19).

These findings allow us to broaden our understanding about the behavior of the virus in the digestive tract and its manifestations, development of preventive measures, and diagnostic and therapeutic strategies (3). In this study, we focused on the demographic, epidemiologic, and clinical characteristics of patients with confirmed SARS-CoV-2 infection at a highly complex institution in Bogota, Colombia, with special emphasis on the presence of gastrointestinal symptoms.

## MATERIALS AND METHODS

### Type of Study

In this quantitative, cross-sectional, analytical, and observational study, we included 72 patients aged >18 years, diagnosed with SARS-CoV-2 infection by analyzing the nasopharyngeal swab samples using Real-Time Reverse Polymerase Chain Reaction (RT-PCR), and who visited the Shaio Clinical Foundation between March 1 and May 31, 2020.

The patients were classified into two groups as follows: presenting and not presenting gastrointestinal symptoms. The definition of gastrointestinal symptoms included the presence of at least one of the following: nausea and /or vomiting, abdominal pain, or diarrhea during patient admission. Anorexia was ruled out as a digestive symptom because it is not an exclusive gastrointestinal symptom. Patients who developed these symptoms during hospitalization or during the progression of their disease were not included to rule out the influence of possible side effects of treatments and other factors. The definition of diarrhea was the presence of more than three bowel movements per day with decreased consistency in patients who did not have a recent history of antibiotic treatment, thus decreasing the chance of *Clostridioides difficile* infection. The severity of respiratory compromise was classified into six degrees in accordance with the recommendation of the Colombian Consensus on Care, Diagnosis, and Management of SARS-CoV-2/COVID-19 infection in Health Care Establishments, published by the Colombian Association of Infectious Diseases and the Institute of Technological Health Assessment of Colombia (1) (Table 1).

**Table 1.** Classification of the severity of respiratory infections

| **Table 1. Classification of the severity of respiratory infections** |  |  |
| --- | --- | --- |
|  | **Degree of severity** | **Definition** |
| **1** | Uncomplicated disease | Local upper respiratory symptoms and may present nonspecific symptoms such as fever, muscle pain, or atypical symptoms in the elderly |
| **2** | Mild pneumonia | Confirmed on chest X-ray and no signs of severity. SaO<sub>2</sub> in ambient air >93%. Consider CURB-65 scale to determine hospitalization |
| **3** | Severe pneumonia | Suspected respiratory infection, failure in one organ, SaO<sub>2</sub> in ambient air, 30 resp/ min |
| **4** | Acute respiratory distress syndrome | Clinical findings, bilateral infiltrated X-ray + oxygenation deficit: mild 200 mmHg < PaO<sub>2</sub>/FiO<sub>2</sub> < 300 mmHg. Moderate: 100 mmHg < PaO<sub>2</sub>/FiO<sub>2</sub> < 200 mmHg. Severe: PaO<sub>2</sub>/FiO<sub>2</sub> < 100 mmHg. If PaO<sub>2</sub> not available consider SaO<sub>2</sub>/FiO<sub>2</sub> |
| **5** | Sepsis | Defined as organic dysfunction that can be identified as a sharp change in the SOFA scale >2 points. Quick SOFA (qSOFA) can identify severe patients with two of the following three clinical variables: Glasgow ≤13, systolic pressure of ≤100 mmHg, and respiratory rate of ≥22 resp/min. Organic insufficiency may occur with the following alterations: acute confusional state, respiratory failure, reduction in diuresis volume, tachycardia, coagulopathy, metabolic acidosis, and increase in lactate level |
| **6** | Septic Shock | Hypotension that persists after resuscitation volume and requires vasopressors to maintain PAM >65 mmHg and lactate >2 mmol/L (18 mg/dL) in the absence of hypovolemia |

### Procedures

Demographic, clinical, and laboratory data were collected from the medical records by specialist physicians and were included in a database designed from a matrix of variables previously reviewed and approved by all authors. The nasopharyngeal swab samples obtained from all patients were transferred to viral medium and processed in laboratories authorized by the National Institute of Health of Colombia to confirm the presence of SARS-CoV-2 by RT-PCR and other viruses including influenza A and B; parainfluenza 1, 2, and 3; adenovirus; and respiratory syncytial virus.

Demographic and clinical data were collected such as comorbidities, presence of respiratory and digestive symptoms at the time of admission, and laboratory results performed within the first 24 h. Results related to the outcome of the disease such as hospitalization time, requirement of admission to the intensive care unit, need for orotracheal intubation, and death were also included.

### Statistical Analysis

For the description of patient characteristics and the variables in general, central trend (average) and dispersion (standard deviation) measures were used in quantitative variables, after comparing the normality in their distribution using Shapiro–Wilk test. If such a situation was not proven, they were described through median and interquartile ranges. Qualitative variables were described and analyzed using absolute frequencies and percentages.

Quantitative variables between the groups were compared using the mean difference Z test on normal-distributed variables. In another case, Wilcoxon range sum test was used. A Z test of difference in proportions was used for qualitative dichotomous variables and Pearson’s χ^2^ test was used for polytomous variables when the expected values in each box were ≥5; otherwise, an exact Fisher’s test was used. The tests were considered statistically significant at an α value of ≤0.05. Statistical analyses were conducted using Stata software (V.15, Stata Corporation, College Station, Texas).

## RESULTS

### Demographic and Epidemiological Characteristics

We included 72 patients with RT-PCR positive for SARS-CoV-2 (34 women and 38 men) with an average age of 47.5 ± 17.7 years; of these, 17 (23.6%) presented at least one of the gastrointestinal symptoms (nausea, vomiting, abdominal pain, and/or diarrhea). Of the 17 patients, 13 (76.47%) presented with diarrhea, five (29.41%) with nausea /vomiting, and five (29.41%) with abdominal pain (Table 2). Diarrhea occurred in 18.06% of all those infected with SARS-CoV-2 at the time of consultation, which was the most common digestive symptom. Patients with gastrointestinal symptoms had an average age of 50 years, with a male–female ratio of 1:1.125. No significant difference was observed in terms of the clinical history or co-existing conditions in those with gastrointestinal symptoms (GIS) vs those without gastrointestinal symptoms (NGIS).

**Table 2.** Demographic and clinical characteristics of patients diagnosed with SARS-CoV-2 infection

|  |  |  |  |
| --- | --- | --- | --- |
| Variables | No gastrointestinal symptoms<br>(n = 55) | Presenting gastrointestinal symptoms<br>(n = 17) | p-Value |
| Demographic variables |  |  |  |
| Gender - (n - %) |  |  |  |
| Female | 25 – 45.45 | 9 – 52.94 | 0.589* |
| Male | 30 – 54.55 | 8 – 47.06 |  |
| Age (years) |  |  |  |
| Median (DE) | 48.23 (18.00) | 45.11 (17.45) | 0.5317** |
| Presence of relevant medical history (n - %) |  |  |  |
| Current smoking (Yes) | 6 -10.91 | 0 – 0.0 | 0.325*** |
| History of smoking (Yes) | 7 – 12.72 | 1 – 15.88 | 0.671*** |
| Arterial hypertension (Yes) | 12 – 21.82 | 5 – 29.41 | 0.527*** |
| Diabetes mellitus (Yes) | 8 – 14.55 | 2 – 11.76 | 1.000*** |
| Chronic kidney disease (Yes) | 1 – 1.82 | 2 – 11.76 | 0.137*** |
| Chronic liver disease (Yes) | 0 – 0.0 | 0 – 0.0 | - |
| Heart failure (Yes) | 2 – 3.64 | 1 – 5.88 | 0.560*** |
| Chronic pulmonary disease (Yes) | 1 – 1.82 | 2 – 11.76 | 0.137*** |
| Pregnancy (Yes) | 0 – 0.0 | 0 – 0.0 | - |
| Immunosuppression (Yes) | 1 – 1.82 | 0 – 0.0 | 1.000*** |
| Cancer (Yes) | 2 – 3.64 | 1 – 5.88 | 0.560*** |
| Hypothyroidism (Yes) | 5 – 9.09 | 2 – 11.76 | 0.665*** |
| Non-gastrointestinal symptoms (n - %) |  |  |  |
| Fever (Yes) | 32 – 58.18 | 11 – 64.71 | 0.632* |
| Anosmia (Yes) | 3 – 5.45 | 2 – 11.76 | 0.586*** |
| Dysgeusia | 6 – 10.91 | 4 – 23.53 | 0.232*** |
| Cough | 48 – 87.27 | 14 – 82.35 | 0.691*** |
| Dyspnea | 32 – 58.18 | 9 – 52.94 | 0.783*** |
| Odynophagia | 23 – 41.82 | 6 – 35.29 | 0.779*** |
| Sputum | 8 – 14.55 | 2 – 11.76 | 1.000*** |
| Dysphonia | 0 – 0.0 | 0 – 0.0 | - |
| Nasal congestion | 1 – 1.82 | 0 – 0.0 | 1.000*** |
| Hemoptysis | 1 – 1.82 | 1 – 5.88 | 0.419** |
| Gastrointestinal symptoms (n - %) |  |  |  |
| Nausea | - | 5 – 29.41 | - |
| Abdominal pain | - | 5 – 29.41 | - |
| Diarrhea | - | 13 – 76.47 | - |
| Abbreviations: SD, Standard Deviation |  |  |  |
| *Differences calculated using Chi-squared test |  |  |  |
| **Differences calculated using Z test of mean difference |  |  |  |
| ***Differences calculated using Fisher's exact test. |  |  |  |

### Clinical Features and Laboratory Results

The most common symptoms in both groups were fever, cough, dyspnea, and odynophagia, but there was no significant difference in their presentation between the GIS and NGIS groups as well as in the other less common presentations including anosmia, dysgeusia, sputum production, nasal congestion, and hemoptysis (Table 2).

Regarding respiratory compromise due to COVID-19 (Table 3), 35 patients in the NGIS group (63.4%) and nine in the GIS group (52.94%) were classified to have uncomplicated disease. Although there was a higher percentage of patients with digestive symptoms in the most severe degrees (severe pneumonia, acute respiratory distress syndrome, sepsis, and septic shock), there were no statistically significant differences (p:0.506). As per the presence or absence of each individual digestive symptom, abdominal pain was the one related to the most severe presentations of COVID-19. This is similar to that reported in a meta-analysis by Kumar et al (20), although there was also no significant difference.

**Table 3.**
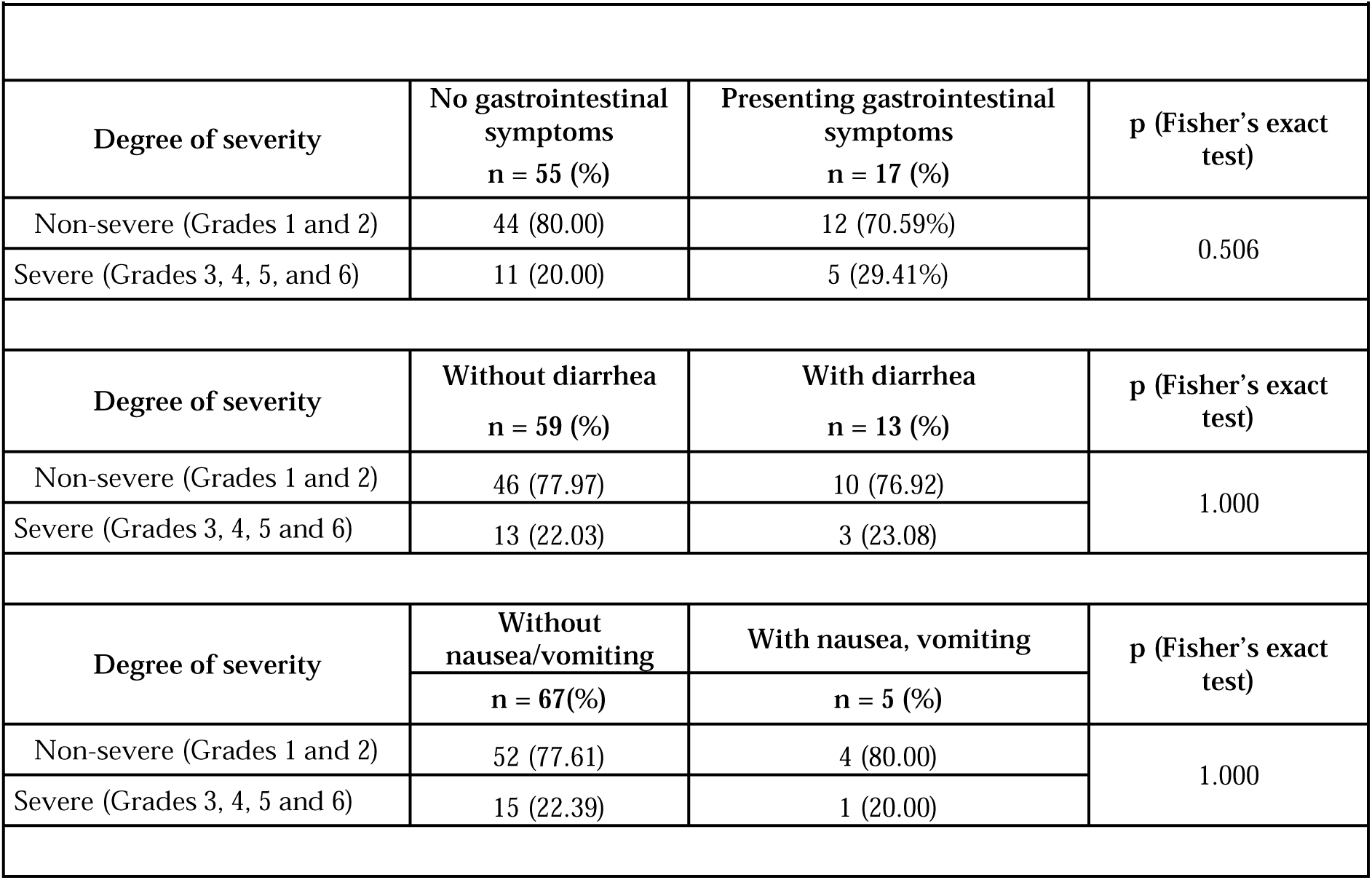
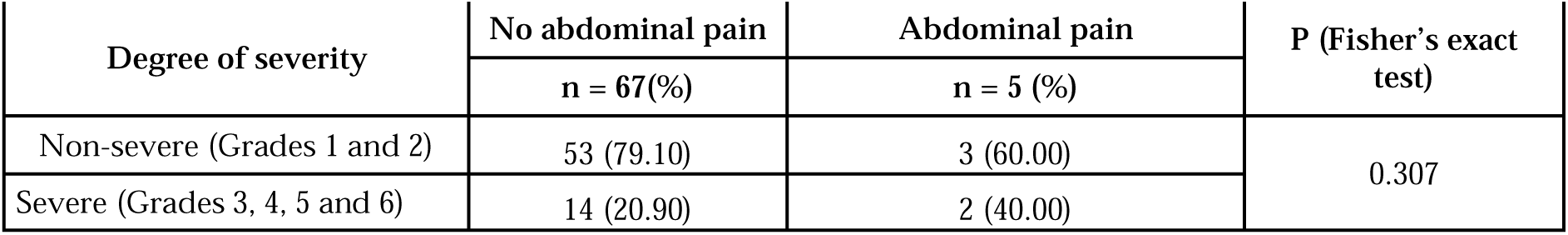
Comparison of respiratory infection severity in patients presenting SARS-CoV-2

Upon comparing the requirement for endotracheal intubation, hospitalization, ICU admission, and fatal outcome between the NGIS and GIS groups, no significant differences were observed in their p-values (0.671, 0.483, 1,000, and 1,000, respectively; Table 4).

**Table 4.** Outcomes in patients with confirmed SARS-CoV-2 infection

| Variables | No gastrointestinal symptoms<br>(n = 55) | Presenting gastrointestinal symptoms<br>(n = 17) | p-Value |
| --- | --- | --- | --- |
| n - % |  |  |  |
| Ventilation | 7 – 12.3 | 1 – 5.88 | 0.671* |
| Hospitalization | 27 – 49.09 | 10 – 58.82 | 0.483** |
| ICU | 8 – 14.55 | 2 – 11.76 | 1.000* |
| Mortality | 4 – 7.27 | 1 – 5.88 | 1.000* |
| *Differences calculated using Fisher's exact test. |  |  |  |
| **Differences calculated using Chi-squared test. |  |  |  |

Regarding the laboratory results, patients in the NGIS presented lower lymphocyte counts (p: 0.044) and higher levels of total bilirubin (p: 0.037) and lactate dehydrogenase (p: 0.014) than those in the SGI group (Table 5). Despite the difference, the lymphocyte count and total bilirubin levels were within normal ranges in both groups. There was no significant difference in the other components of the blood count, renal function, liver enzymes, PCR, ferritin, or D-dimer.

**Table 5.**
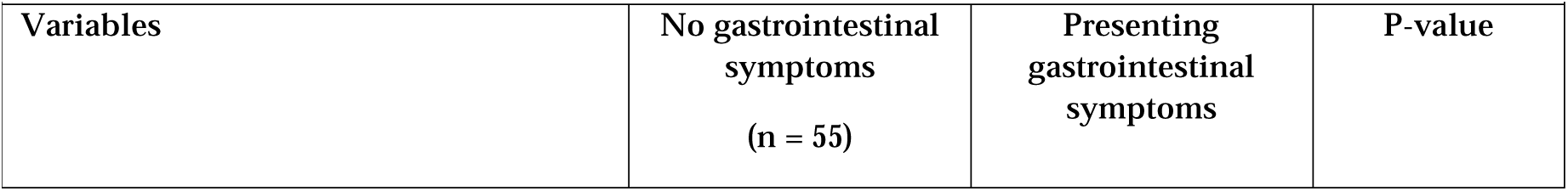
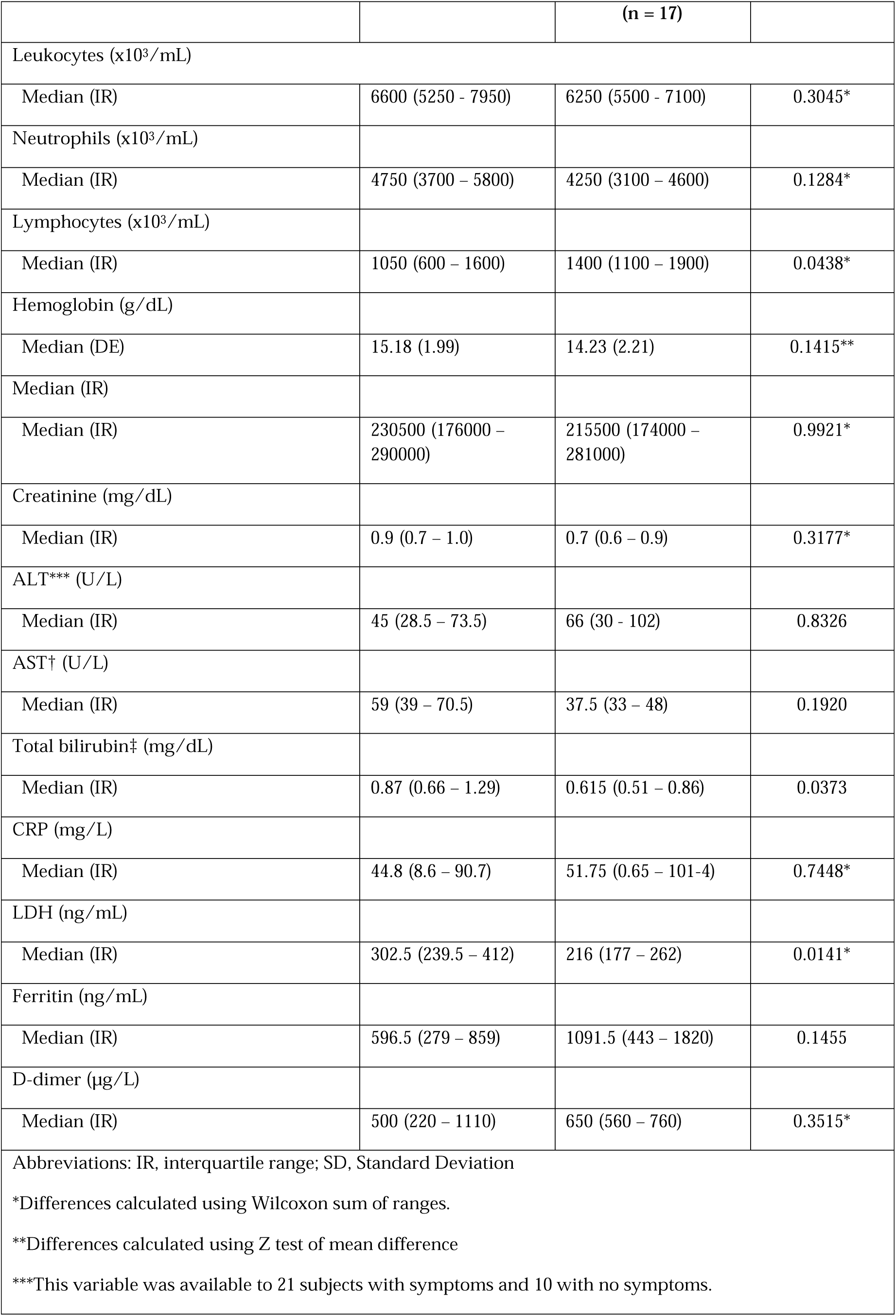
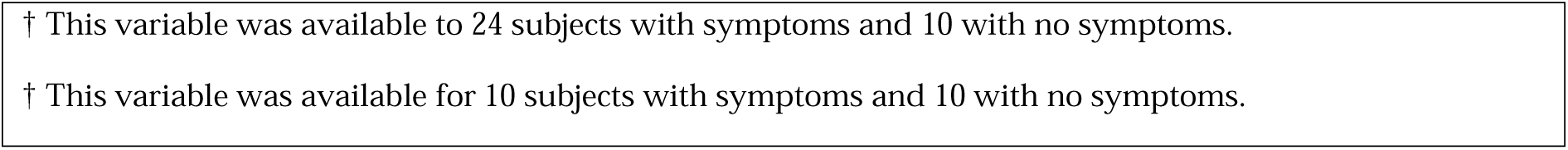
Laboratory results in patients with SARS-CoV-2 infection

| Variables | No gastrointestinal symptoms<br>(n = 55) | Presenting gastrointestinal symptoms | P-value |
| --- | --- | --- | --- |

|  |  | (n = 17) |  |
| --- | --- | --- | --- |
| Leukocytes (x10<sup>3</sup>/mL) |  |  |  |
| Median (IR) | 6600 (5250 - 7950) | 6250 (5500 - 7100) | 0.3045* |
| Neutrophils (x10<sup>3</sup>/mL) |  |  |  |
| Median (IR) | 4750 (3700 – 5800) | 4250 (3100 – 4600) | 0.1284* |
| Lymphocytes (x10<sup>3</sup>/mL) |  |  |  |
| Median (IR) | 1050 (600 – 1600) | 1400 (1100 – 1900) | 0.0438* |
| Hemoglobin (g/dL) |  |  |  |
| Median (DE) | 15.18 (1.99) | 14.23 (2.21) | 0.1415** |
| Median (IR) |  |  |  |
| Median (IR) | 230500 (176000 – 290000) | 215500 (174000 – 281000) | 0.9921* |
| Creatinine (mg/dL) |  |  |  |
| Median (IR) | 0.9 (0.7 – 1.0) | 0.7 (0.6 – 0.9) | 0.3177* |
| ALT*** (U/L) |  |  |  |
| Median (IR) | 45 (28.5 – 73.5) | 66 (30 - 102) | 0.8326 |
| AST† (U/L) |  |  |  |
| Median (IR) | 59 (39 – 70.5) | 37.5 (33 – 48) | 0.1920 |
| Total bilirubin‡ (mg/dL) |  |  |  |
| Median (IR) | 0.87 (0.66 – 1.29) | 0.615 (0.51 – 0.86) | 0.0373 |
| CRP (mg/L) |  |  |  |
| Median (IR) | 44.8 (8.6 – 90.7) | 51.75 (0.65 – 101.4) | 0.7448* |
| LDH (ng/mL) |  |  |  |
| Median (IR) | 302.5 (239.5 – 412) | 216 (177 – 262) | 0.0141* |
| Ferritin (ng/mL) |  |  |  |
| Median (IR) | 596.5 (279 – 859) | 1091.5 (443 – 1820) | 0.1455 |
| D-dimer (µg/L) |  |  |  |
| Median (IR) | 500 (220 – 1110) | 650 (560 – 760) | 0.3515* |
| Abbreviations: IR, interquartile range; SD, Standard Deviation |  |  |  |
| *Differences calculated using Wilcoxon sum of ranges. |  |  |  |
| **Differences calculated using Z test of mean difference |  |  |  |
| ***This variable was available to 21 subjects with symptoms and 10 with no symptoms. |  |  |  |

† This variable was available to 24 subjects with symptoms and 10 with no symptoms.
† This variable was available for 10 subjects with symptoms and 10 with no symptoms.

## DISCUSSION

In this study, 23.6% showed some type of digestive symptom, predominantly diarrhea in 18.05% of the total, which is a finding consistent with several international reports such as that published by Pan et al. (9). However, it is not consistent with the findings of Xiao et al. (8), who reported up to 35.6% patients with diarrhea, or Guan et al. (13), who only found 3.8% patients with this symptom among those positive for SARS-CoV-2.

Several studies, such as those of Wang et al. (10) and Pan et al. (9) in Hubei Province, China, included anorexia as a digestive symptom with a presentation in up to 39.9% patients. However, this symptom was not considered in the present study because it is not specifically related to digestive tract compromise. Although nausea and vomiting are independently described in a majority of international publications, in the present study, they were considered to be a single symptom because they usually occur together. Digestive bleeding has been described as a possible symptom of SARS-CoV-2 infection (14,15) and has been reported in up to 13.7% patients with this condition (8). It was not considered in the analysis of the present study given the significant influence of other external factors and those of patients such as the presence of coagulopathy, acid-peptic disease, use of NSAIDs, and presence of stress ulcers in patients requiring mechanical ventilation in the intensive care unit (which are not the focus of the present study). As a relevant finding in contrast to some previous studies, patients with gastrointestinal symptoms had no significant differences in terms of disease severity, admission to the intensive care unit, or death compared to those who did not have such symptoms. Although lymphocyte count results and LDH and bilirubin levels statistically differed, they were not clinically important because their values were within the normal range.

SARS-CoV-2/COVID 19 infection is a life-threatening disease with significant variability in its clinical presentation and severity of its course. Digestive symptoms are part of the possible range of clinical presentations in patients with this infection, making it necessary to suspect it in patients who, with a potential notion of contagion, present diarrhea, nausea/vomiting, or abdominal pain. The RT-PCR study for SARS-CoV-2 in fecal matter has not yet been standardized in Colombia. However, it would be an important diagnostic tool to identify especially respiratory asymptomatic patients with exclusively digestive symptoms, but with a great capacity for contagion, and thus contribute toward taking public health measures to slow down the rate of the spread of the disease.

Further studies are required to improve the characterization of patients infected with SARS-CoV-2 in local institutions and thus to understand whether there are viral or host factors such as genetic or immunological differences that will allow the development of local strategies aimed at diagnosing and managing such cases.

## Data Availability

FUNDCION CLINICA SHAIO

## Acknowledgements

The authors would like to thank the Ethics and Research Committee of Fundación Clínica Shaio for allowing the data collection.

## Contributors

ACM, RRS, DCZ, and SHT designed the study, collected and analyzed the data, performed the study, and wrote the paper.

JOL designed the study, supervised the entire study process, and wrote the paper.

AMD, JGC, AOH, HIV, JOL, and ESM supervised the entire study process and critically revised the manuscript.

FC designed the study, analyzed the data, and wrote the paper.

## Patient and public involvement

Patients and/or the public were not involved in the design, conduct, reporting, or dissemination plans of this research.

## Patient consent for publication

Not Required

## Ethics approval

The study was approved by the Ethics and Research Committee of the Fundación Clínica Shaio (Minutes 295, April 2020).

